# Psoriasis risk allele function in activated Th1/17 cells with "memory" to antigen exposure

**DOI:** 10.1101/2025.10.22.25338519

**Authors:** Bayazit Yunusbayev, Sergei Ryakhovsky, Radick Altinbaev, Anastasia Kislova, Kseniya Danilko, Liudmila Kraeva, Milyausha Yunusbaeva

**Affiliations:** Institute of Translational Biomedicine, St Petersburg State University, Saint-Petersburg, Russia; Department of Genetics and Biotechnology, St Petersburg State University, Saint-Petersburg, Russia; SCAMT Institute, ITMO University, Saint Petersburg,197101, Russia; Laboratory of Neurophysiology of Learning, Institute of Higher Nervous Activity and Neurophysiology of RAS, Moscow, Russia; Cell Culture Laboratory, Bashkir State Medical University, Ufa, Russia; Laboratory of Medical Bacteriology, Saint-Petersburg Pasteur Institute, Saint-Petersburg, Russia; Laboratory of Functional Genomics, Saint-Petersburg State Phthisiopulmonology Research Institute, Saint-Petersburg, Russia

## Abstract

Most causal variants for complex diseases are expected to affect gene regulation in a cell- and context-specific manner. Hence, identification of such dynamically functioning variants requires functional readouts in disease-relevant tissues and context. In this study, we prioritized causal variants for psoriasis by enhancing functional annotations from disease-relevant cells. We demonstrate that disease-relevant immune cells, unlike most body tissues, possess functional annotations matching candidate causal SNPs. Specifically, we identified an eQTL rs4672505 affecting *B3GNT2* gene expression only in Th1/Th17 cells with a memory phenotype, i.e., antigen-experienced cells. This eQTL also matched an enhancer chromatin mark exclusive to memory T cells and absent in other tissues. For this eQTL rs4672505, the risk allele A reduces *B3GNT2* gene expression. B3GNT2 deficiency in murine models reduces the glycosylation of the CD28 co-receptor and increases T cell activation upon antigen stimulation. Notably, memory T cells also require CD28 co-stimulation, which occurs only when antigen-presenting cells encounter an infection. Thus, our findings suggest that the risk allele likely increases co-stimulation of memory Th1/Th17 cells by their cognate antigens under infection. This putative mechanism aligns with the observation that psoriasis can manifest, exacerbate, and recur after infection.

## Introduction

Most causal variants (∼90%) for inflammatory diseases are expected to be non-coding and affect disease risk through gene regulation (1) and splicing (2). For non-coding variants, traditional challenges of fine-mapping, such as strong linkage disequilibrium (LD), are further complicated by the difficulty in identifying the relevant cell type and effector gene that mediates function (3). This is because non-coding variants contribute to disease by affecting different stages of gene regulation and splicing (2). Gene regulation is dynamic, often in unknown cell populations and specific physiological contexts (4). This is also true for psoriasis, for which dozens of genome-wide association studies (GWAS) have suggested nearly 100 risk loci; however, so far, only a few risk loci have been fine-mapped and studied functionally (5).

Often, statistical fine-mapping highlights multiple candidate SNPs in LD, and further prioritization is based on finding overlaps with functional annotations, such as genomic sites matching with expression quantitative trait loci (from here onwards, simply eQTLs), open- chromatin regions, and other annotations predicting regulatory function (6). However, a major problem is that for any given disease, disease-specific functional annotations (e.g., eQTL) can be missing in the widely used databases of functional annotations. For example, popular resources, such as The Genotype-Tissue Expression (GTEx) (7), The Encyclopedia of DNA Elements (ENCODE) (8), and NIH Roadmap Epigenomics (ROADMAP) (9), represent only a fraction of the known functional variants in the human genome, as described in tissues from healthy donors or patients with select disorders. In the meantime, causal variants are non- coding and expected to function dynamically in disease-specific tissues under a particular physiological context that is often unknown (10). Thus, even with state-of-the-art fine-mapping tools that integrate large databases for functional annotations, such as GTEx, most disease-associated regulatory SNPs may be missing. Therefore, to achieve systematic functional mapping, one must manually collect/curate functional genomic annotations tailored to the disease in focus, along with relevant immune cell types and physiological states.

In this study, we focused on fine-mapping known risk loci for psoriasis, for which previous large-scale studies had unresolved causal SNPs and effector genes. Previous studies have nominated effector genes for psoriasis risk loci, initially using the nearest gene approach and more recently employing a transcriptome-wide association study (TWAS) (5,11–13). Fine-mapping causal SNPs involves predicting the effector gene, the "gene for which the product is predicted to mediate the effect of a genetically associated variant on a disease" (3). As we show in this study, promising candidates for causal SNPs and effector genes can be missed even when using powerful tools that rely on functional annotations from standard tissues not relevant for disease. In this study, to increase fine-mapping success, we considered a wide range of cells and tissues and maximized functional annotations from primary immune cells relevant to disease (14) (15). We avoided functional readouts from cells with perturbed genetic and epigenetic genomic landscapes, such as immortalized cell lines or induced pluripotent stem cells (iPS cells). We also paid attention to the fact that psoriasis, like other autoimmune diseases, can be triggered by exposure to infections (16,17), and immunopathology involves immune cells in an activated but not naive state. Hence, we tried to add functional annotations where immune cells were activated, challenged by diverse microbial ligands, or had memory of previous antigen exposure. Specifically, we expanded the standard eQTL data set from 48 GTEx tissues (7,18) with eQTLs from 23 immune cells and states published elsewhere (18–22). By incorporating more datasets on immune cells into our analyses, we maximized the discovery of regulatory SNPs (eQTLs) that demonstrated regulatory function depending on the state of the immune cells—naive, activated, or memory state. In this way, we tested our hypothesis that part of the missing psoriasis-associated causal variants regulates immune-related genes when cells respond to infection or danger signals.

## Results

### Statistical fine-mapping

In this study, we employed the SUSIE fine-mapping approach to prioritize causal SNPs within 64 psoriasis risk loci (out of a total of 67) previously reported in (13) (S1 Table). These risk loci were previously analyzed in (5) using meta-analysis and functional fine-mapping. We start by highlighting commonly encountered outcomes in statistical fine-mapping to make several points essential for practice. First, SNPs in strong LD to the lead SNP can sometimes be causal variants. Hence, focus on the lead SNP for functional study, as was done previously (23), can be potentially misleading if not supported by statistical fine-mapping. Secondly, strong LD in some risk loci makes statistical fine-mapping inefficient, and functional annotations or experiments are needed to prioritize functional SNPs.

We begin by demonstrating an example, where statistical fine-mapping was sufficient to prioritize the most likely causal variant with good evidence, i.e., with high posterior probability of being causal, PP ≥ 0.95 (from here onward, posterior probability (PP) or probability). In Fig. 1a, the lead SNP rs8016947 (indicated by a red diamond) has multiple adjacent variants with comparable p-values ("GWAS track", Fig 1a). In this psoriasis locus, statistical fine-mapping ("SUSIE track", Fig 1a) prioritized the lead SNP rs8016947 as the most likely (indicated by a red-green circle) causal variant (PP ≥ 0.95). This kind of desired fine-mapping outcome, however, represents a rare example (∼9%, 6 out of 64 loci), where statistical fine-mapping alone was sufficient to prioritize the causal variant with good evidence (high PP ≥ 0.95). More often (∼36%, 23 out of 64 loci), however, the degree of certainty in fine-mapping, as indicated by the posterior probability, is relatively low (well below 0.95). Usually, the top-prioritized SNP has only marginally higher PP than nearby SNPs (Fig 1b). For psoriasis, we inferred 23 such risk loci, where lead SNPs scored somewhat higher evidence than nearby SNPs, but had posterior probabilities well below 0.95. For instance, in Fig 1b, the ’GWAS’ track highlights a lead SNP, rs11249215, in the previously established risk locus with index SNP rs7536201. The lead SNP rs11249215 (red diamond), which falls outside the *RUNX3* coding sequence, is flanked by dozens of SNPs in strong LD (r^2^ ≥ 0.8) that have comparable p-values. Our statistical fine-mapping with SUSIE (Fig 1b, ’SUSIE’ track) suggested that this non-coding lead SNP rs11249215 had only marginally higher PP (PP=0.257) than flanking SNPs (PP varied from 0.03 to 0.05).

**Fig 1.**
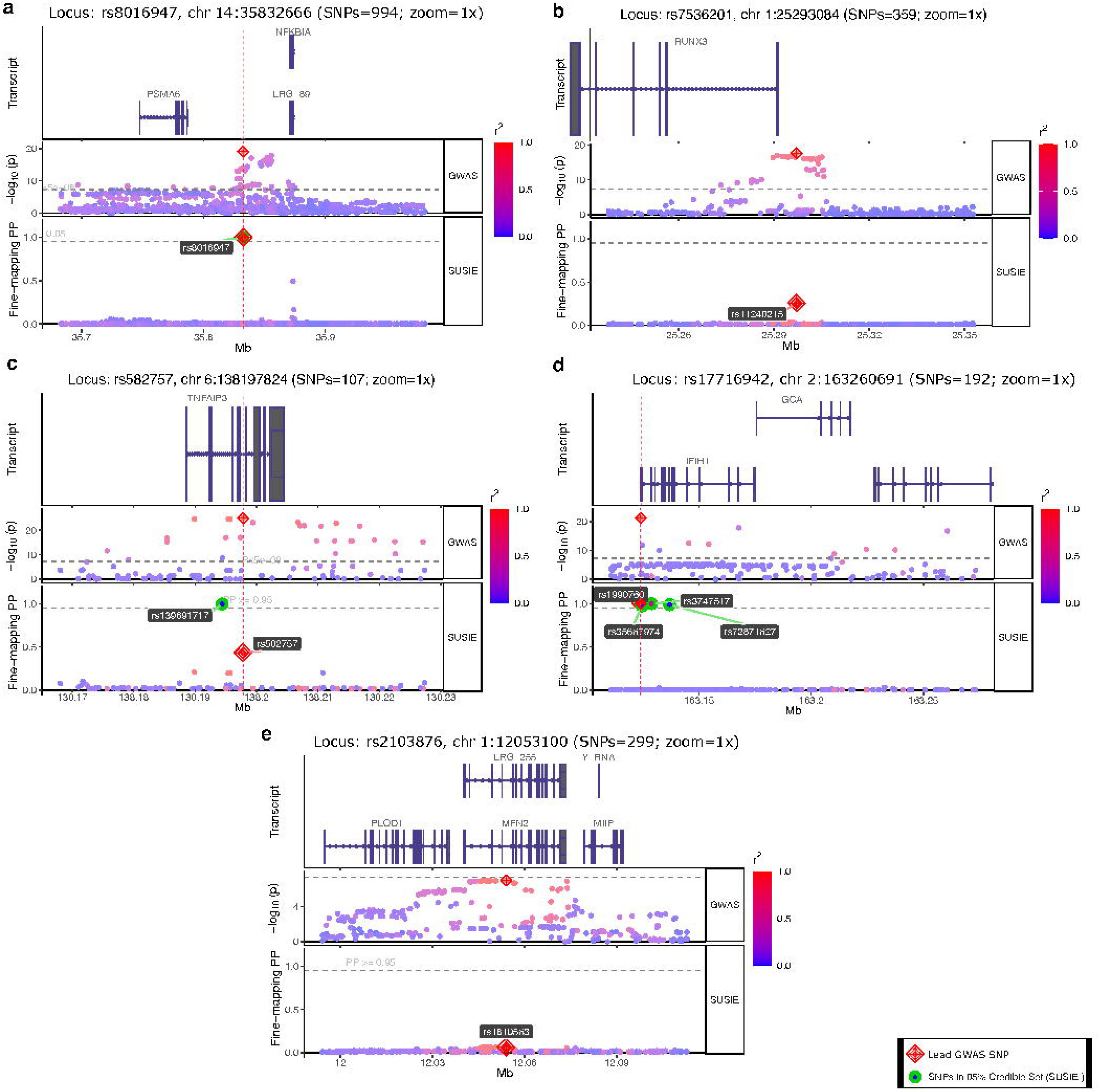
Five types of statistical fine-mapping outcomes. Figure panels a, b, c, d, and e depict annotation tracks for five psoriasis risk loci. For each risk locus, the upper track depicts the gene transcripts in the locus, the middle ’GWAS’ track depicts log-transformed GWAS p-values for SNPs, and the bottom ’SUSIE’ track represents fine-mapping results with posterior probabilities. The green circle indicates SNPs within 95% SUSIE Credible Set, i.e., predicted causal variants with strong support (PP ≥ 0.95). Red diamond indicates the lead GWAS SNP. (a) Statistical fine-mapping outcome: the GWAS lead SNP was prioritized as the most likely causal variant with strong evidence (PP ≥ 0.95). (b) The top-prioritized causal variant had only marginally higher PP than nearby SNPs. Panel c. Non-lead SNP had strong evidence of being causal (PP ≥ 0.95). Panel d. Multiple SNPs had strong evidence (PP ≥ 0.95) of being causal. Panel e. Strong LD rendered multiple causal SNP configurations equally possible, each with weak support. Statistical fine-mapping was ineffective.

P-values reported by GWAS measure the probability of chance finding, but not the likelihood of causality. For that reason, the lead SNP with the lowest p-value does not necessarily represent the causal SNP. For example, a non-causal SNP can be inferred as the lead SNP when it is in LD with two independent causal variants. In this scenario, a non-causal SNP combines the effects of two SNPs and may achieve a stronger association than the marginal associations of the two independent true SNPs (24). Yet, for psoriasis, earlier studies took lead SNPs from GWAS to design functional follow-up analyses (23). Our statistical fine-mapping revealed seven risk loci for psoriasis (7 out of 64 risk loci), where non-lead SNPs achieved comparable or higher probabilities of being causal. For example, Fig 1c illustrates GWAS SNPs in the rs582757 locus, scattered around the *TNFAIP3* gene transcript. There were multiple genome-wide significant SNPs in high LD, with the lead SNP rs582757 ("GWAS track", Fig 1c). According to our fine-mapping analysis, one of the non-lead SNPs, rs139691717 (indicated by a green circle), has strong evidence of being causal (PP = 0.996). In contrast, the lead SNP, rs582757, had much less support (PP = 0.433) for being causal. We also encountered four loci (4 out of 65) where the lead SNP and one or more nearby SNPs were jointly inferred to be causal. For example, in two cases, the lead SNP and three nearby SNPs all had strong evidence (PP ≥ 0.95) of being causal (’SUSIE’ track, Fig 1d), while in two other loci, the lead SNP and nearby SNPs all had identical but low PP (PP < 0.95) (Not shown).

Finally, strong linkage disequilibrium often (∼37% of loci, 24 out of 65) renders fine-mapping ineffective. With strong LD (r2 ≥ 0.8), many causal SNP configurations are equally possible, and many SNPs have individually small posterior probabilities. Fig 1e showcases this frequent fine-mapping outcome. In this example, the lead SNP, rs1810563, in the previously established risk locus (with index SNP rs2103876), received weak evidence (PP well below 0.2) of being causal, similar to other linked SNPs in the vicinity (Fig 1e).

### Colocalizing GWAS SNPs with eQTLs from 48 GTEx tissues and 23 immune cells

Like an earlier study (5), our statistical fine-mapping with SUSIE prioritized an extensive collection of probable causal SNPs within 64 risk loci (established in (13)) (S1 Table). Most risk loci are presumed to influence disease through an impact on gene expression (25). To link our prioritized SNPs with nearby gene expression, we performed colocalization analysis with eQTL markers from different tissues. In total, we run colocalization for cis-acting eQTLs affecting gene expression in 48 human body tissues/cells (7,18) and 23 immune cell types and states (naïve, activated, and memory cells) (18–22) (S2 Table). Altogether, we colocalized 194 eQTLs with PP4D≥D0.8 at a 10% false discovery rate (FDR) across 69 (out of 71 analyzed) tissues and cell types (S3 Table). Of these, 39 were in high LD (r² ≥ 0.8) (Fig 2) (S4 Table), with 13 index SNPs in the previously established 64 risk loci (S1 Table).

**Fig 2.**
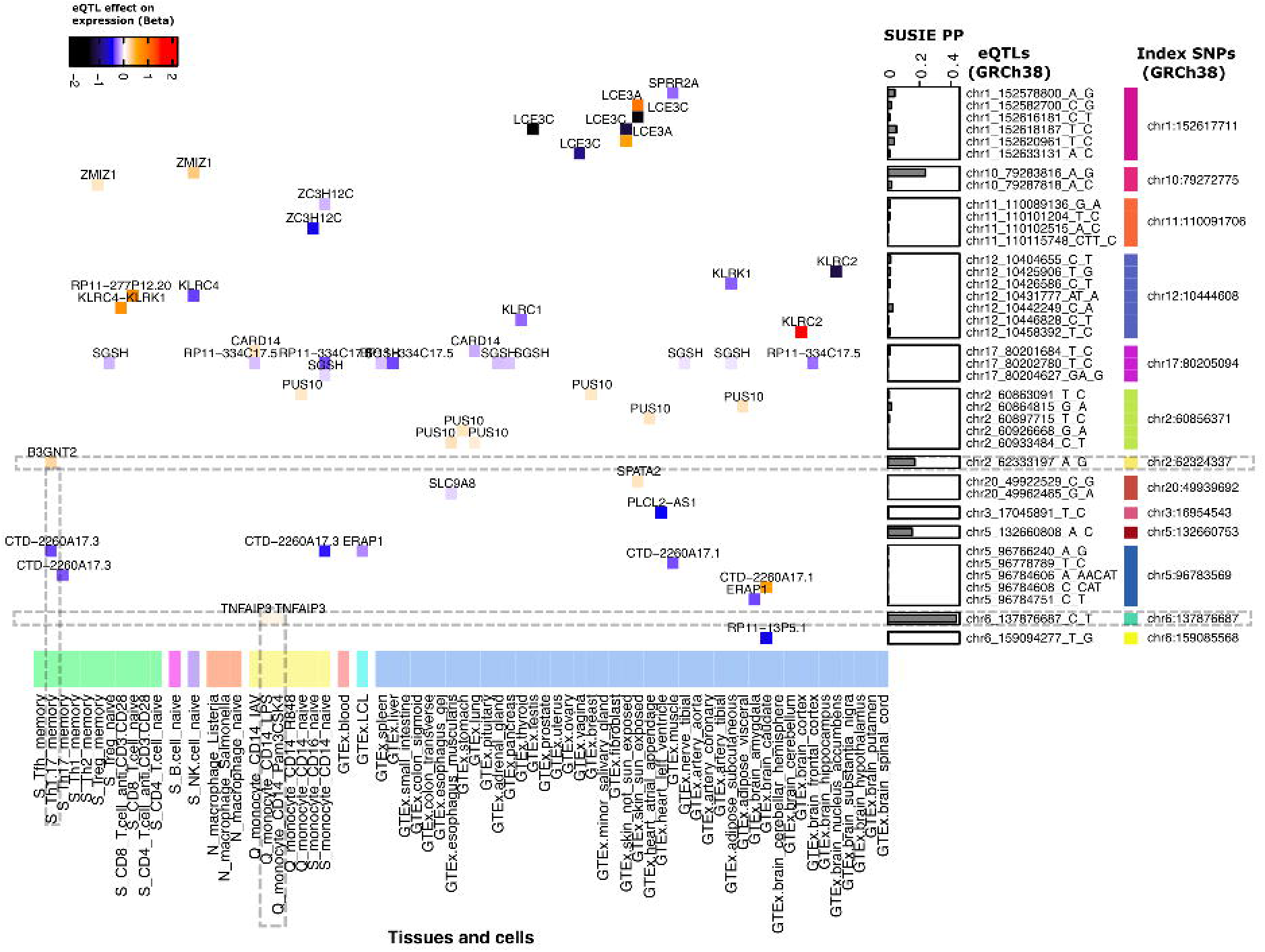
Expression QTLs colocalized with GWAS SNPs. Heatmap cells, the color-filled rectangles, represent colocalized eQTLs with effector gene names. Each eQTL has a line with the genomic position (GRCh38) and a column with the cell/tissue, where it was identified in the original eQTL mapping study. For each colocalized eQTL, the rectangle color indicates whether its effect allele increases (from white to red) or decreases (from white to black) the effector gene expression. For instance, in the upper left corner, C alelle for the eQTL chr10_79287818_A_C increases ZMIZ gene expression in T regulatory cells with memory phenotype (’S_Treg_memory’). Note that this eQTL chr10_79287818_A_C has another nearby eQTL chr10_79283816_A_G that also affects ZMIZ gene expression, but in naive NK cells ’S_NK.cell_naive’, and this eQTL has a higher posterior probability of being causal (PP) based on SUSIE fine-mapping. Both eQTLs were in strong LD with the index SNP chr10:79272775.

### Prioritized causal SNP with putative regulatory function in effector/memory T cells

We used SUSIE-computed posterior probabilities to determine if any of the 39 eQTLs showed evidence of being causal within the 13 risk loci. Most of the 39 eQTLs (within 9 out of the total 13 risk loci) lacked strong evidence of being causal. None of them stood out with a high probability (PP ≥ 0.95) to prioritize. Instead, most of them, on average, had relatively small PP (PP ≤ 0.1) (See "SUSIE PP" bar plots in Fig 2). Nevertheless, our extended panel of immune cells (altogether 23) with different phenotypes (naive, activated, memory) allowed us to discover and prioritize two likely functional SNPs among probable causal SNPs within two risk loci (with index SNP rs10865331 and rs582757, respectively) (S1 Table). These putative functional SNPs, the two eQTLs (indicated by horizontal dashed lines in Fig 2), could have been missed if we had analysed only GTEx tissues, as many other studies do. Furthermore, we highlighted these two eQTLs because experimental tests would be easier to establish for them, as they function in a single cell type. Also, it would be easier to find supporting (but independent) functional annotations in specific cell types and then test their absence in other cells and tissues. Taking into account these practical considerations, we focused on one of the discovered eQTLs, rs4672505 (indicated as chr2_62333197_A_G and marked by a horizontal dashed line in Fig 2), which was present only in memory Th1/17 cells (highlighted by a vertical dashed line in Fig 2). Since this eQTL was absent in other T cell types and GTEx tissues, we reasoned that its function must be specific to this memory Th1/17 population. These memory Th1/17 cells (denoted as ’S_Th1.17_memory’ in Fig 2) were defined by a surface marker set (CD3+/CD4+/CD25low/CD45RA-/CD127high/CCR3-/CCR4+/CCR6+) in the original study (19) and represent a mixture of T helper 1 and T helper 17 cells with memory phenotype (Th1 and Th17 cells from here onward).

### Active chromatin state specific to effector/memory T cells

The memory phenotype for these Th1 and Th17 cells indicates that these T cells were previously activated by their cognate antigen (26). We, therefore, hypothesized that our eQTL site, rs4672505, likely overlaps a regulatory element that opens when naive CD4+ T cells transition into a memory state by remodeling their chromatin into an active state. Our hypothesis assumes that all other cells and tissues must lack this active chromatin state at our eQTL site. Moreover, this eQTL site must also be repressed at all stages of T cell ontogeny, i.e., before naive T cells turn into memory cells. To test this working hypothesis, we screened active (e.g., active transcription start sites (TTS) and enhancers) and inactive chromatin states (e.g., repressed PolyComb) in different tissues and immune cells available in the ROADMAP dataset (S5 Table) (9). Specifically, we examined chromatin peaks around our eQTL site rs4672505 and for the entire risk locus. The ROADMAP dataset allowed us to screen chromatin marks in all the immune cells representing major steps in the immune cell ontogeny: starting with common hematopoietic stem cells, and ending with major derived cell fates, such as myeloid cells (monocytes and neutrophils), lymphoid cells (T cells, B cells), and Natural Killer cells. We did not find clear chromatin peaks indicating active chromatin states at the queried eQTL site, neither in hematopoietic stem cells nor among major derived branches, such as myeloid cells (monocytes and neutrophils), and most lymphoid cells (B-cells and some T cells, different from CD4+ helper cells (Fig 3a). Reassuringly, we also did not find active chromatin states in non-immune tissues (S1 Fig).

**Fig 3.**
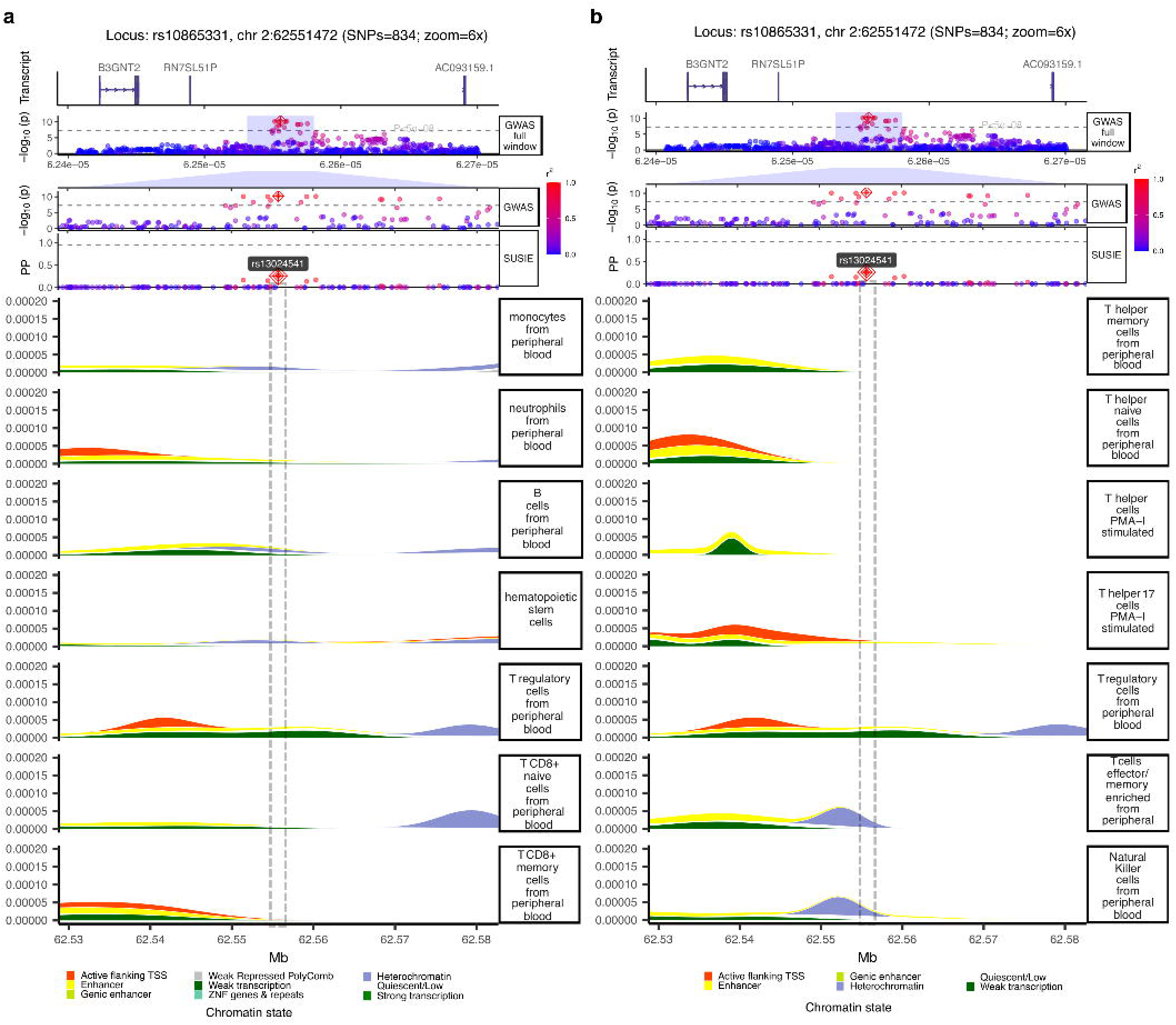
Chromatin states from different cell types combined with fine-mapping results for the risk locus rs10865331. Our target eQTL, rs4672505, is very close to the lead SNP rs13024541 at the centre of the psoriasis risk locus with index SNP rs10865331. Figure tracks from top to bottom depict gene transcript annotations ’Transcript’ for the risk locus, GWAS results, SUSIE-based statistical fine-mapping, and chromatin states from different cell types.

Consistent with our eQTL-based hypothesis (eQTL unique to memory T cells, highlighted in Fig 2), we found clear cell-specific chromatin peaks at our focal eQTL rs4672505 in memory T cells (Fig 3b). We found two chromatin states that overlapped each other; one associated with an enhancer (active state) and another with a heterochromatin (inactive) mark, respectively (Enhancer peak is behind heterochromatin peak in Fig 3b, see also S2 Fig). We also found similar chromatin peaks among Natural Killer cells (NK cells) (Fig 3b). However, these chromatin peaks can not be reliably attributed to NK cells because of the uncertainty regarding the cell type purity. NK cells in the ROADMAP project were defined by gating CD56- positive cells (See link to the source data in S6 Table) (9). It is known that the CD56 marker is also expressed by many immune cells, including alpha beta T cells, gamma delta T cells, dendritic cells, and monocytes (27). Hence, the chromatin marks observed among the so-called "NK cells" can be a readout from any of these CD56-positive cells. We, therefore, conclude that the identified active and inactive chromatin marks at our eQTL site rs4672505 can be reliably attributed only to memory T cells, which were defined as ’CD4+_CD25int_CD127+_Tmem_Primary_Cells’ in the ROADMAP project. Finally, one can notice that several T cell varieties (T CD8+, T helper, T regulatory) have transcription-associated active chromatin state (’Active flanking TSS’) around the *B3GNT2* gene, the effector gene for our eQTL (Fig 3). This transcription signal widespread among T cells agrees with the fact that transcription-associated states tend to be more constitutive across related cell types. In contrast, enhancer-associated states are highly tissue-specific (9).

In summary, our results suggest that observed immune-cell-specific eQTLs from the eQTL Catalogue and chromatin marks from the ROADMAP provide two independent lines of evidence that our fine-mapped SNP, rs4672505, represents a regulatory variant at genomic position GRCh38.2:62333197:A/G. Given eQTL findings, allele G is associated with the higher expression of the *B3GNT2* gene in memory in Th1/17 cells (Fig 2). In contrast, the alternative A allele is associated with a lower *B3GNT2* gene expression, and this allele is associated with psoriasis risk. According to the literature, B3GNT2 protein deficiency reduces post-translational modification of immune cell surface receptors and makes T cells more reactive. Specifically, B3GNT2*-*deficient mice have a lower polylactosamine decoration of CD28 and CD19 co-stimulatory molecules on T helper cells. Lower polylactosamine makes these immune cells more sensitive to induction by antigen, and results in a stronger T cell proliferation (28).

## Discussion

### Statistical fine-mapping alone is not enough to resolve promising causal variants

We show that statistical fine-mapping alone delivers only a few clear candidates for experimental follow-up, which agrees with current observations in the field. According to the literature, for complex diseases, only a few risk loci have been characterized mechanistically (6). For example, for psoriasis, the latest fine-mapping study prioritized 14 candidate variants with highly probable regulatory functions (5). Dand et al. reported a list of candidate causal SNPs, LD blocks, and multiple genes falling into 109 independent association signals (summarized in Table 10 in the original study)(5). Reported credible sets with causal SNPs had an average size of 74 kb and contained dozens of highly correlated SNPs, in strong LD. Poor fine-mapping resolution is explained by strong linkage disequilibrium, which is rooted in the unique demographic history of the extant human populations (29). Namely, most human populations stem from a recent (∼50 000 years ago) strong bottleneck event from Africa (30).

In our study, for most of the risk loci, statistical fine-mapping was also not enough to narrow down individual candidates for causal variants (Fig 1). For example, for our focal risk locus tagged by index SNP rs10865331 ("GWAS full window" and "GWAS" tracks in Fig 3a), we observed several strongly linked candidate SNP (rs13024541 with the highest PP) with comparable SUSIE PP ("SUSIE" track in Fig 3a), which hindered causal SNP prioritisation. For the same risk locus, a previous study used genotypes from European and trans-ethnic cohorts and managed to narrow down (95% Bayesian Credible Interval) 9 and 11 SNPs, respectively (13).

Strong LD often prevents fine-mapping in the study population, which usually (∼90%) consists of patients with European ancestry (31). In such cases, one can achieve further progress by adding trans-ethnic cohorts, if available (13). In practice, however, multi-ancestry GWAS data (32) are rare, and the application of multi-ancestry fine-mapping is rare. More often, further prioritization is achieved by functional annotation-informed fine-mapping (33). i.e., by adding functional annotations. This approach critically depends on the availability of functional readouts from disease-relevant tissues (33). Moreover, our eQTL-based findings suggest that the discovery of functional variants potentially related to disease can also be dependent on the physiological state of the implicated cell.

### Systematic fine-mapping assisted by functional annotations

To prioritize causal variants among linked SNPs in our risk locus of interest (with index SNP rs10865331, defined as chr2:62551472 in (13), we added functional annotations. We sought SNPs with a function in immune cells relevant to psoriasis pathogenesis. By adding 23 immune cell types on top of the 49 GTEx tissues (S2 Table), we found a novel eQTL that was present only in memory Th1/17 cells. This helped us to prioritize a potentially functional SNP with a regulatory effect on the nearby *B3GNT2* gene expression restricted to memory Th1/17 cells. The affected *B3GNT2* gene was more distal to the eQTL (rs4672505) compared to other more proximal transcripts, annotated in this region ("Transcript" track in Fig 3a). Starting with the earliest post-GWAS studies, the *B3GNT2* gene was suspected as the effector gene, first based on proximity to the index SNP (34). Currently, it is widely accepted that hand-picking a proximal gene as an effector gene can be misleading unless supported by a rigorous fine-mapping procedure (3). Recently, the TWAS approach helped to include the *B3GNT2* gene into the list of candidate effector genes (5). Unfortunately, the TWAS approach has known pitfalls. This approach "does not allow for any conclusions to be drawn about the relationship of the phenotype with the gene expression" (35). Finally, for our risk locus of interest (with index SNP rs10865331), a recent study inferred a different effector gene (than *B3GNT2)*, the *COMMD1* gene, approximately 435 kb upstream of the locus (23). This study used a capture Hi-C (CHi- C) approach to find a long-range interaction between the index SNP rs10865331 and the *COMMD1*, but not *B3GNT2* (23). This inference is problematic for two reasons. Firstly, index SNP rs10865331 was picked as a candidate causal variant without performing statistical fine-mapping. In our study, we deliberately reported SUSIE fine-mapping outcomes to emphasize that the lead (and index) SNP does not always represent the causal variant (Fig 1). This is also likely true for our focal risk locus ("SUSIE" track in Fig 3a), where a non-lead SNP, rs13024541, was a more promising candidate. Secondly, the authors did CHi-C interaction experiments using immortalized keratinocytes and CD8 T cells (23). Immortalized cell lines have strongly disturbed regulatory circuitry. Specifically, they have substantial differences in chromatin accessibility, nucleosome positioning, histone modifications, and DNA methylation (36) compared to primary cells. Therefore, capture Hi-C readouts reported in (23) may reflect an altered regulatory landscape in immortalized cell lines. It is not clear whether the observed long-range loops to the distal *COMMD1* gene (23) are present in the primary cells. In sum, previous studies suggested effector genes for the rs10865331 locus without following or omitting key steps required for systematic fine-mapping. Moreover, earlier studies did not identify the cell type involved in the disease-associated SNP function, molecular mechanism (promoter, enhancer, or transcription factor (TF) binding site), and whether the risk allele increased or decreased the effector gene expression. In this study, we inferred the cell and context-specific eQTL that links the risk SNP allele effect with the likely effector gene and cell type. These are key parameters to set up experiments and study how effector genes contribute to the pathogenesis.

### The place and timing of the putative causal SNP function

Our findings suggest that psoriasis patients who carry the risk allele at the rs4672505 site likely have lower *B3GNT2* gene expression in memory T helper cells. Lower *B3GNT2* expression can reduce CD28 co-receptor decoration on memory T helper cells. Memory T helper cells, like naive cells, require CD28 co-stimulation to activate when they recognize their cognate antigen. The CD28 co-stimulation can happen only when antigen-presenting cells (APC) express the B7 receptor upon infection (37). These details are important to understanding how the *B3GNT2-*related risk factor could be mechanistically involved in the pathogenesis. First, lower CD28 receptor decoration can make T helper memory cells more sensitive to activation by their cognate antigen. In other words, this genetic predisposition factor may contribute to pathogenesis by increasing the propensity to react to a specific antigen.

Furthermore, cognate antigen is not sufficient to activate memory T helper cells. Antigens recognized by T cells must be presented together with the CD28 co-stimulation via B7 receptor expression on APC. We emphasize that expression of B7 proteins on APC requires microbial exposure (37). Taken together, one can hypothesize that memory T helper cells in psoriasis patients must encounter self-antigens in the context of infection, which correlates with known facts about psoriasis. Indeed, psoriasis often manifests, exacerbates, or reappears when patients encounter infections (38,39). Thus, if our proposed effector gene is validated, it would be interesting to know whether the proposed phenotype, higher reactivity to antigens, requires microbial exposure.

### Broader implications for the current post-GWAS efforts

It is now widely recognized that causal variants for immune-mediated diseases may function dynamically (10,40,41). For example, they can occur in dynamic regulatory elements that "stay quiescent" until immune cells activate, for example, in response to cell-specific environmental cues/triggers (10). This dynamic nature of regulation agrees with psoriasis often manifesting after environmental stress, such as infection (42). Indeed, our fine-mapped SNP (rs4672505, 2:62560332:A:G) was registered as an eQTL in Th1 and Th17 cells, specifically in cells with a memory phenotype, that is, cells that were once activated by an antigen. Hence, our colocalized eQTL would have been missed if functional annotations came from broadly defined immune cell types, such as CD+ T cells, not studied under different functional states — namely, antigen-activated T cells with a memory phenotype relevant for autoimmunity. Our findings exemplify one of the early ideas that deciphering disease genetics would involve tedious work on finding disease-relevant cells and modelling disease-relevant context (40). Our results support the idea that filling the missing link between disease risk loci and function can not be achieved only by adding more readouts (such as pQTLs, splice variants, chromatin marks, etc) from the same tissues. Instead, more effort is needed to experiment with disease-relevant tissues under disease-relevant physiological states.

### Implications for the follow-up experiments

Our findings offer a hypothesis for directing experimental research - information about the cell type and molecular phenotype to test in vitro or in vivo. Specifically, one has to focus on primary Th1 and Th17 cell populations from patients, but not reprogrammed or otherwise manipulated cell cultures to recapitulate the unique chromatin landscape around the causal SNP. Cell populations from risk allele carriers and non-carriers can be tested for *B3GNT2* gene expression and polylactosamine decoration of the CD28 receptors. The associated pathogenic phenotype, according to our hypothesis, must be differential sensitivity to activation by cognate antigen. Cognate autoantigens can be based, for example, on known protein epitopes (43). Taken together, our findings suggest that experimental follow-up should take care of all the details - focus on primary cell type, memory phenotype, and cognate antigen challenge in the context of infection. If confirmed, polylactosamine decoration, memory T cells, or even infections in patients can be a potential therapeutic target.

## Materials and Methods

### GWAS dataset and statistical fine-mapping

To carry out genetic fine-mapping for psoriasis risk loci, we used GWAS summary statistics for 9 010 555 SNPs published earlier (13). The GWAS summary statistics were obtained from the NHGRI-EBI Catalog https://www.ebi.ac.uk/gwas using the study identifier GCST90019016. To organize candidate SNPs around established psoriasis risk loci, we used 64 index SNPs reported in the Table S5 in the original study (13) (S1 Table). Before fine-mapping, we performed quality control (QC) and harmonization using the MungeSumstats R package (44). Altogether, 7 791 492 SNPs were retained for downstream analyses after QC and harmonization. To carry out fine-mapping, we integrated summary statistics with the linkage disequilibrium (LD) data from a reference population of European ancestry (EUR) in the 1000 Genomes project phase 3 (45). Statistical fine-mapping step was performed on each locus separately with susieR (24) using the echolocatoR() pipeline (46). For each locus, we assumed a maximum of five causal SNPs and ran fine-mapping using variable genomic window sizes (30 kb, 60 kb, 150 kb, 250 kb, and 450 kb). We chose the window size that best captured the SNPs in LD, with the lead SNP. For each locus, fine-mapping inferred the posterior probability (PP) of each SNP being causal. Depending on the local distribution of SNP posterior probabilities, we defined three fine-mapping outcomes: a) One or more SNPs have strong evidence (PP ≥ 0.95) of being causal, b) one or more SNPs demonstrate higher PP of being causal than nearby SNPs, but probabilities are low (PP < 0.95) c) multiple SNPs are strongly linked and demonstrate similar but low PP of being causal.

### Colocalization of GWAS associations with eQTLs

The GWAS SNP coordinates for psoriasis were lifted to the GRCh38 reference genome using CrossMap (47) to match with the eQTL variant coordinates. We performed statistical colocalization of psoriasis GWAS SNPs with eQTLs obtained from the eQTL Catalogue (48), which stores uniformly standardized eQTL summary data from published sources. For colocalization, we used QTLs from 48 GTEx human body tissues (7,18) and 23 immune cell types and states (naïve, activated, and memory cells) (18–22). While the eQTL Catalogue featured over 35 immune-related cells, we selected only 23 of them (S2 Table) by excluding reprogrammed iPS cells (20) and cells genotyped on microarrays (18). The major problem with the use of eQTLs from iPS-reprogrammed cells is that such cells retain a residual epigenetic memory of the source somatic cells and have de novo epigenetic aberrations (49). To run colocalization analyses, we used the R package coloc (v.3.1) (50). The P-value distributions for eQTL analyses were post-processed using the qvalue R package (version 2.28.076) to define the p-value threshold corresponding to a 10% false discovery rate (FDR). Co-localization between QTL and psoriasis SNP was considered significant if the target hypothesis achieved strong evidence, as indicated by a posterior probability (PP4D≥D0.8). Here, PP4 stands for the posterior probability of hypothesis 4 when there is a shared SNP association with disease and eQTL. Colocalization analysis computes support (in terms of posterior probability) for all possible configurations (hypotheses), such as, for example, PP1 for hypothesis 1 - SNP association with disease, but not with eQTL, or PP3 for hypothesis 3 - SNP association with disease and eQTL, but there are two independent SNPs. For the total set of 7 791 492 GWAS SNPs with summary statistics, we found 194 eQTL-tissue pairs with PP4D≥D0.8 at 10% FDR (S3 Table). For our analyses, we retained only 39 colocalized eQTLs that fell within one of the 64 established psoriasis risk loci. Specifically, eQTLs were retained if they were in strong LD with one of the lead SNPs from 64 risk loci.

### Overlap with the ROADMAP chromatin states associated with regulatory elements

To find overlaps between colocalized eQTLs and regulatory elements, we used chromatin state annotations from the ROADMAP project (9). We analyzed chromatin states in genomic windows that contained our eQTL of interest and affected genes. In doing so, we first explored chromatin states in the same cell type where the focal eQTL was discovered. We next tested whether our cell-specific eQTL matched similar chromatin states in other cell types and tissues. ROADMAP chromatin states for each cell/tissue type were retrieved and plotted using the echoplot() function in the echolocatoR R package suite (46). Each cell type was accessed using Epigenome ID (EID) (S5 Table), as listed in the original study (9). For example, chromatin states for "Primary hematopoietic stem cells" were retrieved using "E035" Epigenome ID.

## Data Availability

The data underlying the results presented in the study are available from the Figshare open access repository, at https://doi.org/10.6084/m9.figshare.27038266.v1.

## Supporting information

**S1 Fig. Chromatin states from different body tissues combined with fine-mapping results for the risk locus rs10865331.** Our target eQTL, rs4672505, is very close to the lead SNP rs13024541 at the center of the psoriasis risk locus with index SNP rs10865331. Figure tracks from top to bottom depict gene transcript annotations ’Transcript’ for the risk locus, GWAS results, SUSIE-based statistical fine-mapping, and chromatin states from different cell types.

S2 Fig. Chromatin peaks in T effector/memory cells within the risk locus rs10865331.

**S1 Table. Genomic coordinates for psoriasis risk loci.** Psoriasis risk loci were defined based on index SNPs reported by (13), in Table S5 of the original study. Specifically, we used SNP IDs from the "ID" column.

**S2 Table. Expression QTL datasets used for colocalization with psoriasis candidate risk SNPs.** eQTL summary data for each tissue/cell type were retrieved from the eQTL Catalogue project (https://www.ebi.ac.uk/eqtl/). eQTL Catalogue stores uniformly processed gene expression and splicing QTLs from published studies.

**S3 Table. Expression QTLs colocalized with psoriasis GWAS SNPs.** Matrix columns correspond to eQTLs from different tissue/cell types. Matrix rows correspond to psoriasis GWAS SNPs that showed strong evidence (Posterior Probability, PP4 ≥ 0.8) for colocalization with a given eQTL at a given row. Each matrix cell contains a gene name for the affected gene and a Beta-value. Beta-value has a positive or negative value depending on whether the eQTL effect allele increases or decreases gene expression. For example, consider the cell entry, "*ZMIZ1* +0.290125", at the intersection of the first line, "chr10_79283816_A_G", and the column "Schmiedel_2018*NK-cell_naive_ge". Here, the G allele for eQTL chr10_79283816_A_G increases the expression of the *ZMIZ1* gene in naive NK-cells (Schmiedel_2018*NK- cell_naive_ge) proportional to Beta = +0.290125 relative to the alternative A allele.

**S4 Table. Colocalized expression QTLs in strong LD with psoriasis GWAS signals.** Colocalized eQTLs were tested for linkage disequilibrium with index SNPs for known psoriasis risk loci using pairwise r2. Altogether 39 eQTLs were found to be in strong LD (r2 ≥ 0.8) and were retained for analyses.

S5 Table. **С**hromatin state annotations from ROADMAP screened in this study.

Cells and tissues for which chromatin peaks were screened using echoplot::plot_locus() function in the echolocatoR package (46). Each cell and tissue was retrieved using unique epigenome IDs.

S6 Table. Chromatin state annotations as presented in Fig 3.

List of cells and tissues from ROADMAP presented in Fig 3.

