## Supplementary Information for "Psoriasis risk allele function in activated Th1/17 cells with "memory" to antigen exposure"

**Bayazit Yunusbayev^1,2^*****, Sergei Ryakhovsky^1,3^ , Radick Altinbaev^4^ , Anastasia Kislova^1^ , Kseniya Danilko^5^ , Liudmila Kraeva^6^ , Milyausha Yunusbaeva^1,7^**

^1^Institute of Translational Biomedicine, St Petersburg State University,

Saint-Petersburg, 199034, Russia

^2^Department of Genetics and Biotechnology, St Petersburg State University,

Saint-Petersburg, 199034, Russia

^3^SCAMT Institute, ITMO University, Saint Petersburg,197101, Russia

^4^Institute of Higher Nervous Activity and Neurophysiology of RAS,

Laboratory of Neurophysiology of Learning, Moscow, 117485, Russia

^5^Bashkir State Medical University, Cell Culture Laboratory, Ufa, 450008, Russia

^6^Saint-Petersburg Pasteur Institute, Laboratory of Medical Bacteriology,

Saint-Petersburg, 197101, Russia

^7^Saint-Petersburg State Phthisiopulmonology Research Institute, Saint-Petersburg, 191036, Russia

Supplementary Figures

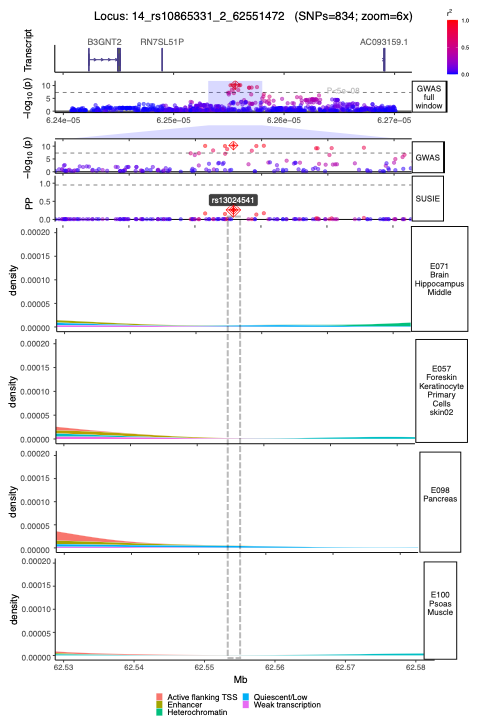

Fig. S1 | **Chromatin states from different body tissues combined with fine-mapping results for the risk locus rs10865331**.

Our target eQTL, rs4672505, is very close to the lead SNP rs13024541 at the center of the psoriasis risk locus with index SNP rs10865331. Figure tracks from top to bottom depict gene transcript annotations 'Transcript' for the risk locus, GWAS results, SUSIE-based statistical fine-mapping, and chromatin states from different cell types.

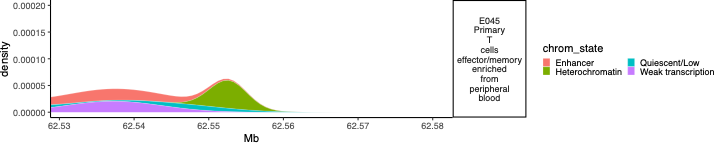

Fig. S2 | **Chromatin peaks in T effector/memory cells within the risk locus rs10865331.**
